# Vitamin D supplementation improves the prognosis of patients with colorectal cancer liver metastases

**DOI:** 10.1101/2022.11.02.22281865

**Authors:** Miran Rada, Lucyna Krzywon, Audrey Kapelanski-Lamoureux, Diane Kim, Stephanie Petrillo, Anthoula Lazaris, Peter Metrakos

## Abstract

Colorectal cancer liver metastasis (CRCLM) is one of the deadliest cancers. CRCLM tumours have two distinct histopathological growth patterns (HGPs) including desmoplastic HGP (DHGP) and replacement HGP (RHGP). The DHGP tumours are angiogenic, while their RHGP counterparts are vessel co-opting. The patients with DHGP tumours have a better response to anti-angiogenic agents and chemotherapy, as well as the prognosis. To determine the influence of vitamin D supplementation in CRCLM, we analyzed the HGPs and the 5-year OS of CRCLM patients (n=106). Interestingly, we found an inverse correlation between vitamin D supplementation and the presence of RHGP tumours in CRCLM patients. Additionally, the 5-year OS of the patients that administered vitamin D was significantly higher. The cancer cells in RHGP lesions are characterized by direct contact with the hepatocytes, and this phenomenon enhances the motility of the cancer cells and facilitates their infiltration through liver parenchyma to co-opt the pre-existing vessels. Significantly, our in vitro data demonstrated the downregulation of motility markers in the co-cultured cancer cells with hepatocytes upon exposure to vitamin D. Altogether, this study highlights the role of vitamin D in CRCLM and provides a rationale to investigate the contribution of vitamin D supplementation to the prognosis of CRCLM patients.

## Introduction

Colorectal cancer (CRC) ranked as the second most lethal cancer worldwide^1^, which accounts for approximately 10% of cancer-related death among men and women^2^. Metastasis is the main cause of death in CRC patients, with approximately 50% of the patients developing liver metastases (LM) during the course of their disease^3^. Surgical removal of the tumour is the only chance to cure patients with colorectal cancer liver metastasis (CRCLM)^4^. However, the ratio of eligible CRCLM patients for hepatic resection is as low as 15-20%^5^. The rest of the patients are referred to chemotherapy combined with targeted therapies such as anti-angiogenic agents (e.g. Bevacizumab)^6,7^. However, the treatment of CRCLM patients is challenging due to the acquired resistance resulting in treatment failure and cancer recurrence^8^.

CRCLM tumours have two major histopathological growth patterns (HGPs) including desmoplastic HGP (DHGP) and replacement HGP (RHGP)^8–11^. In DHGP, the cancer cells are separated from the liver parenchyma via desmoplastic ring^8^. However, the cancer cells of RHGP tumours are in direct contact with liver parenchyma due to the lack of desmoplastic ring^8,12,13^. Additionally, the vascularization in the DHGP lesions is angiogenic, while the RHGP lesions utilize vessel co-option vascularization^8^. In vessel co-opting CRCLM, the cancer cells infiltrate through liver parenchyma and co-opt the pre-existing vessels to obtain the blood supply^8,14^. Of note, vessel co-option is associated with acquired resistance against anti-angiogenic agents and chemotherapy in CRCLM^8^ and other types of cancers, such as hepatocellular carcinoma (HCC) and glioblastoma^15^.

Vitamin D is a secosteroid hormone that is predominately metabolized in the liver and it plays an essential role in calcium homeostasis and bone health^16^. There is increasing importance in regulating vitamin D levels for cancer management. Vitamin D deficiency has been associated with the development of cancer^16,17^. Many laboratory and animal studies linked vitamin D supplementation with lower levels of carcinogenesis and tumour progression, as well as enhanced tumour response to anti-cancer therapies^18^. Chandler et al.^19^ have performed a clinical trial with 25,871 patients to determine the influence of vitamin D supplementation on developing advanced (metastatic or fatal) cancer. Accordingly, vitamin D supplementation reduced the risk of developing advanced cancer^19^. Moreover, their investigation suggested that the effect of vitamin D was more significant among the patients that had normal body mass index (BMI), while no significant impact was found among overweight (BMI 25-29.9) or obese patients (BMI≥30)^19^.

Angiogenesis is the formation of new blood vessels, which plays a pathological role in tumour metastasis^11,20^. Previous investigations suggested vitamin D as both anti-angiogenic and pro-angiogenic agent in cancer^21^. Various studies confirmed the anti-angiogenic effect of vitamin D in various types of cancers^22,23^. However, little is known about the pro-angiogenic effect of vitamin D, particularly in tumours. In vitro study by Dehghani et al.^24^ proposed vitamin D as an inhibitor of apoptosis in endothelial cells. Of note, the endothelial cells are essential for tumour angiogenesis^25^. Moreover, exposing endothelial colony-forming cells to vitamin D significantly induced the expression of vascular endothelial growth factor (VEGF)^26^, a key mediator of tumour angiogenesis^25^. These data suggest that the effect of vitamin D on tumour angiogenesis is controversial and require further investigations.

Our previous publications showed a positive correlation between cancer cell motility and the development of vessel co-option in CRCLM in vivo^12,27,28^, which is mediated by upregulation of runt related transcription factor-1 and actin-related protein 2/3 complex (RUNX1-ARP2/3) pathway^29^. Interestingly, vitamin D has been shown as a negative regulator of cancer cell motility in different types of cancers, such as thyroid^30^, breast^31^, ovarian^32^, and colorectal cancer^33^. The impact of vitamin D on cancer cell motility is yet to be investigated in CRCLM. The role of vitamin D in suppressing non-angiogenic tumour vascularization is poorly understood. Recently, Bajbouj et al.^34^ suggested vitamin D as an inhibitor of non-angiogenic tumours in breast cancers, specifically the tumours that utilize vasculogenic mimicry. However, the effect of vitamin D on vessel co-option is still unknown.

In this manuscript, we found an inverse correlation between the presence of RHGP tumours and vitamin D supplementation in CRCLM patients. Moreover, vitamin D administration significantly improved the 5-year OS of CRCLM patients. Furthermore, our in vitro data suggested that vitamin D attenuates the expression of motility and Epithelial–mesenchymal transition (EMT) markers in cancer cells, which play a crucial role in the generation of vessel co-option tumours in CRCLM.

## Materials and methods

### Clinical data and patient samples

The study was conducted in accordance with the guidelines approved by McGill University Health Centre Institutional Review Board (IRB). Informed consent was obtained from all patients through the McGill University Health Centre (MUHC) Liver Disease Biobank. This study was performed on 84 CRCLM patients who were referred to McGill University Health Centre (MUHC) in Montreal. For patient characteristics see Table 1.

**Table 1.**
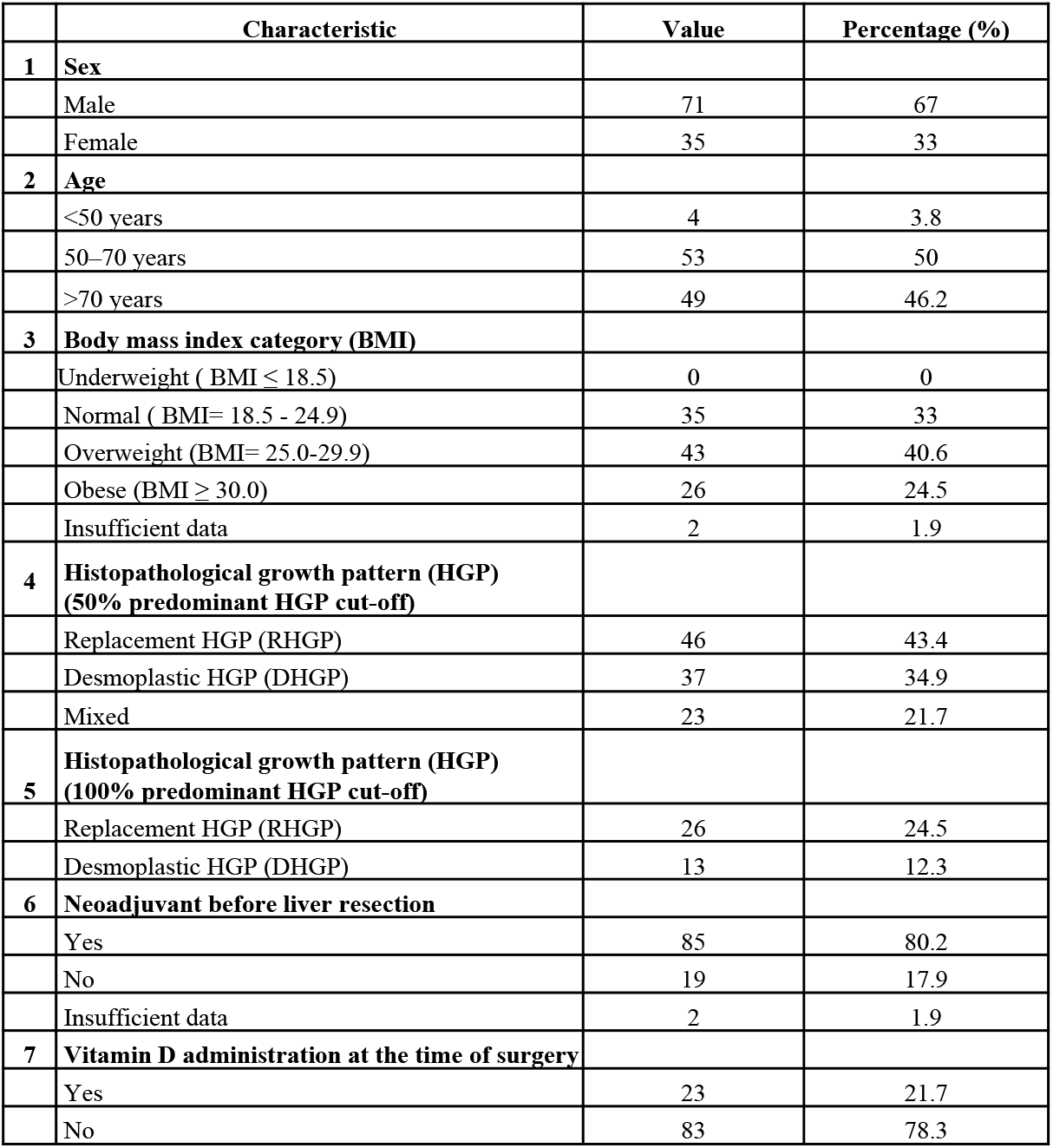
Demographic baseline of CRCLM patients.

|  | Characteristic | Value | Percentage (%) |
| --- | --- | --- | --- |
| 1 | Sex |  |  |
|  | Male | 71 | 67 |
|  | Female | 35 | 33 |
| 2 | Age |  |  |
|  | <50 years | 4 | 3.8 |
|  | 50–70 years | 53 | 50 |
|  | >70 years | 49 | 46.2 |
| 3 | Body mass index category (BMI) |  |  |
|  | Underweight ( BMI ≤ 18.5) | 0 | 0 |
|  | Normal ( BMI= 18.5 - 24.9) | 35 | 33 |
|  | Overweight (BMI= 25.0-29.9) | 43 | 40.6 |
|  | Obese (BMI ≥ 30.0) | 26 | 24.5 |
|  | Insufficient data | 2 | 1.9 |
| 4 | Histopathological growth pattern (HGP)<br>(50% predominant HGP cut-off) |  |  |
|  | Replacement HGP (RHGP) | 46 | 43.4 |
|  | Desmoplastic HGP (DHGP) | 37 | 34.9 |
|  | Mixed | 23 | 21.7 |
| 5 | Histopathological growth pattern (HGP)<br>(100% predominant HGP cut-off) |  |  |
|  | Replacement HGP (RHGP) | 26 | 24.5 |
|  | Desmoplastic HGP (DHGP) | 13 | 12.3 |
| 6 | Neoadjuvant before liver resection |  |  |
|  | Yes | 85 | 80.2 |
|  | No | 19 | 17.9 |
|  | Insufficient data | 2 | 1.9 |
| 7 | Vitamin D administration at the time of surgery |  |  |
|  | Yes | 23 | 21.7 |
|  | No | 83 | 78.3 |

### Kaplan-Meier estimates of overall survival

Overall survival estimates were calculated from the date of diagnosis of liver metastases to the date of death or to the date of the last follow-up.

### Cell culture

Human colorectal cancer (HT29) and hepatocyte (IHH) cells were cultured^9,10^ in DMEM (Wisent Inc., #319-005-CL) supplemented with 10% FBS (Wisent Inc., #085-150) and 1× penicillin/streptomycin (Wisent Inc., 450-201-EL). All cells were cultured at 37 °C with 5% CO2.

### Immunofluorescence staining

Immunofluorescence staining was performed for co-cultured HT29 cancer cells that co-cultured with IHH hepatocytes as described in previous publications^9,13,35^.

### Statistical analysis

Statistical analysis was performed using GraphPad Prism software version 7.0 (GraphPad Software, La Jolla, CA, USA) software. Unpaired Student’s t-test was applied to compare the means of two groups. The association between the two categorical groups was assessed with the Chi-square test. For the overall survival data, the Log-Rank test was used to determine the statistical significance. P-values of <0.05 were considered to be significant. Data presented as mean ± standard deviation.

## Results

### Vitamin D supplementation induces the generation of angiogenic tumours and improves the prognosis

For the clinical aspects of vitamin D supplementation in CRCLM patients, we collected and analyzed a retrospective cohort of 106 patients (summarized in Table 1) who underwent CRCLM resection at the McGill University Health Centre, Montreal, Quebec, Canada. We stratified the patients into three groups based on the HGPs (50% predominant HGPs cut-off) of their tumours including RHGP, DHGP and mixed (RHGP, DHGP and/or pushing HGP). We found a better survival rate for the patients with DHGP tumours than those who had RHGP or mixed tumours (Figure 1a). Similar results were found when we applied further stratification and compared the patients with 100% DHGP or 100% RHGP tumours (Figure 1b).

**Figure 1.**
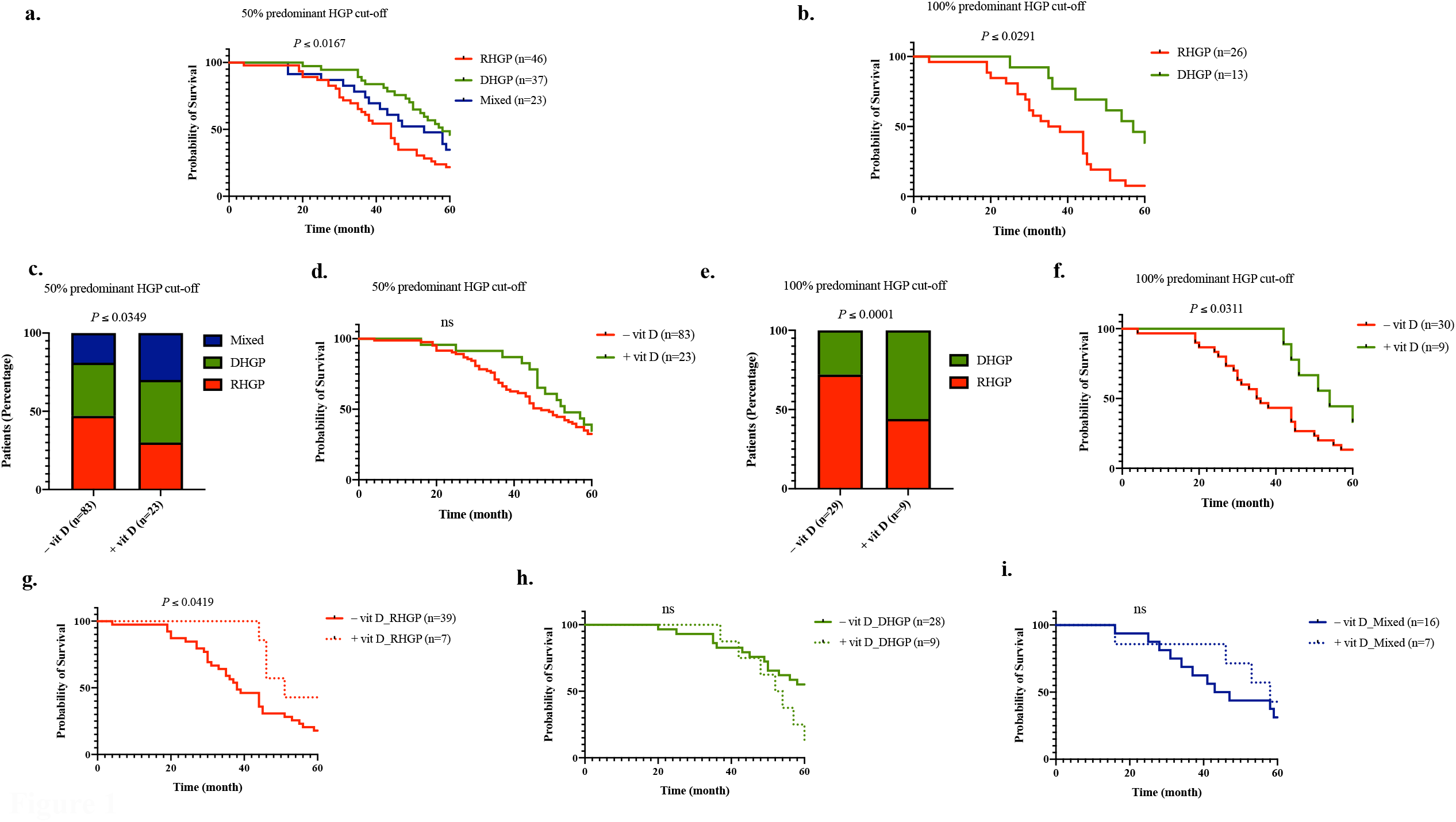
Vitamin D supplementation the 5-year overall survival of CRCLM patients. **a**. and **b**. Kaplan–Meier survival analysis shows 5-year OS of 106 patients with CRCLM. **c**. Represents the percentage of patients with vessel co-option (RHGP), angiogenic (DHGP) or mixed (RHGP and DHGP) lesions according to the supplementation of vitamin D. The HGP of the CRCLM tumours were scored based on 50% cut-off predominant HGP. **d**. Kaplan–Meier survival analysis shows the effect of vitamin D supplementation on 5-year OS of CRCLM patients. The HGP of the patients was scored based on 50% cut-off predominant HGP. **e**. Shows the percentage of patients with vessel co-option (RHGP) and angiogenic (DHGP) lesions upon vitamin D supplementation. The HGP of the CRCLM tumours was scored based on 100% cut-off predominant HGP. **f**. Kaplan– Meier survival analysis shows the association between vitamin D supplementation and 5-year OS of CRCLM patients. The HGP of the CRCLM tumours was scored based on 100% cut-off predominant HGP. **g-i**. Kaplan–Meier survival analysis shows the influence of vitamin D supplementation in CRCLM patients with either RHGP, DHGP, or mixed tumours. The HGP of the patients was scored based on 50% cut-off predominant HGP.

We then evaluated whether vitamin D supplementation would correlate with the presence of co-opting RHGP tumours. Surprisingly, we found a lower percentage of patients with RHGP tumours in the subgroup of the patients that used vitamin D (Figure 1c). Importantly, the survival rate of the patients that used vitamin D was also higher than the rest of the patients (Figure 1d). Indeed, the influence of vitamin D supplementation was more prominent when we analyzed only the patients that had either 100% DHGP or 100% RHGP tumours (Figures e and f). We further analyzed our cohort and assessed the 5-year overall survival (OS) in the patients with RHGP tumours upon their administration of vitamin D. As shown in Figure 1g, vitamin D supplementation was significantly associated with a better 5-years OS in the patients with RHGP tumours. However, no correlation was found when we examined the influence of vitamin D on 5-year OS in the patients with DHGP or mixed tumours (Figures 1h and 1i).

To further evaluate the effect of vitamin D supplementation on CRCLM, we compared the number and diameter of the tumours of the CRCLM patients that used vitamin D supplementation to the rest of the patients. We noticed a dramatic reduction in the number and diameter of the tumours upon vitamin D supplementation (Figures 2a and b). Collectively, these data suggest a positive effect of vitamin D on the prognosis of CRCLM patients, which is possibly incited by its effect on the vascularization, number and diameter of the tumours.

**Figure 2.**
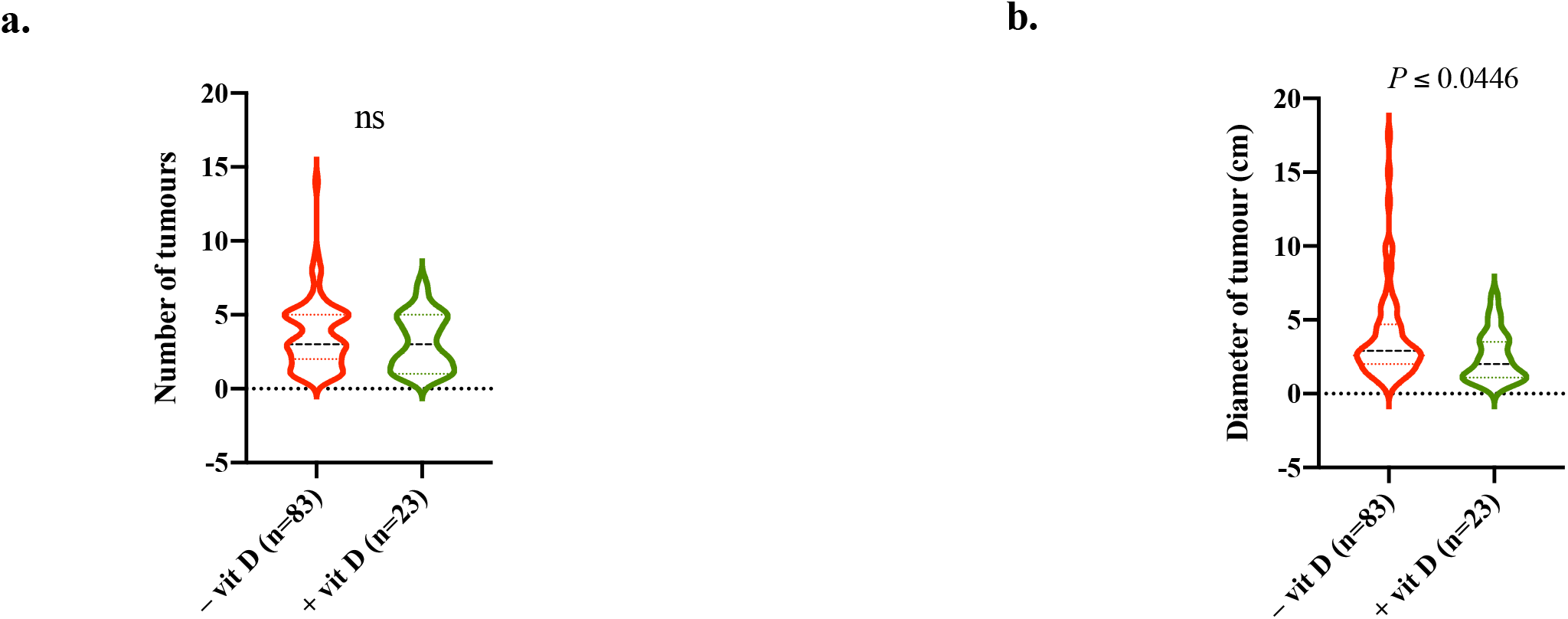
Vitamin D supplementation associated with smaller tumours in CRCLM. **a**. and **b**. Show the number and diameter of the tumours, respectively, in CRCLM patients in the presence or absence of vitamin D supplementation.

### The benefit of vitamin D is correlated with the BMI of the patients

Various studies suggested the association between BMI and cancer mortality^36–38^. Accordingly, overweight patients have the lowest odds of cancer death than the rest. On the other hand, it has been reported that the benefit of vitamin D supplementation is significantly higher in cancer patients with normal weight than in overweight and obese patients^19^. Therefore, we decided to evaluate the effect of vitamin D in CRCLM based on the BMI of the patients. The patients that administrated vitamin D were stratified into two groups based on their BMI including normal weight (n=8) and overweight/obese (n=15) patients. Intriguingly, the percentage of patients with RHGP tumours was significantly lower in the patients with normal weight compared to overweight/obese patients (Figure 3a). The overweight/obese patients were similar to normal wright patients when we compared the number of tumours (Figure 3b). However, the normal-weight patients had smaller tumours than their counterparts (Figure 3c). Most importantly, the overweight/obese patients had significantly lower 5-year OS than normal-weight patients. Taken together, these data suggest vitamin D supplementation is less effective in overweight/obese CRCLM patients than in their normal-weight counterparts.

**Figure 3.**
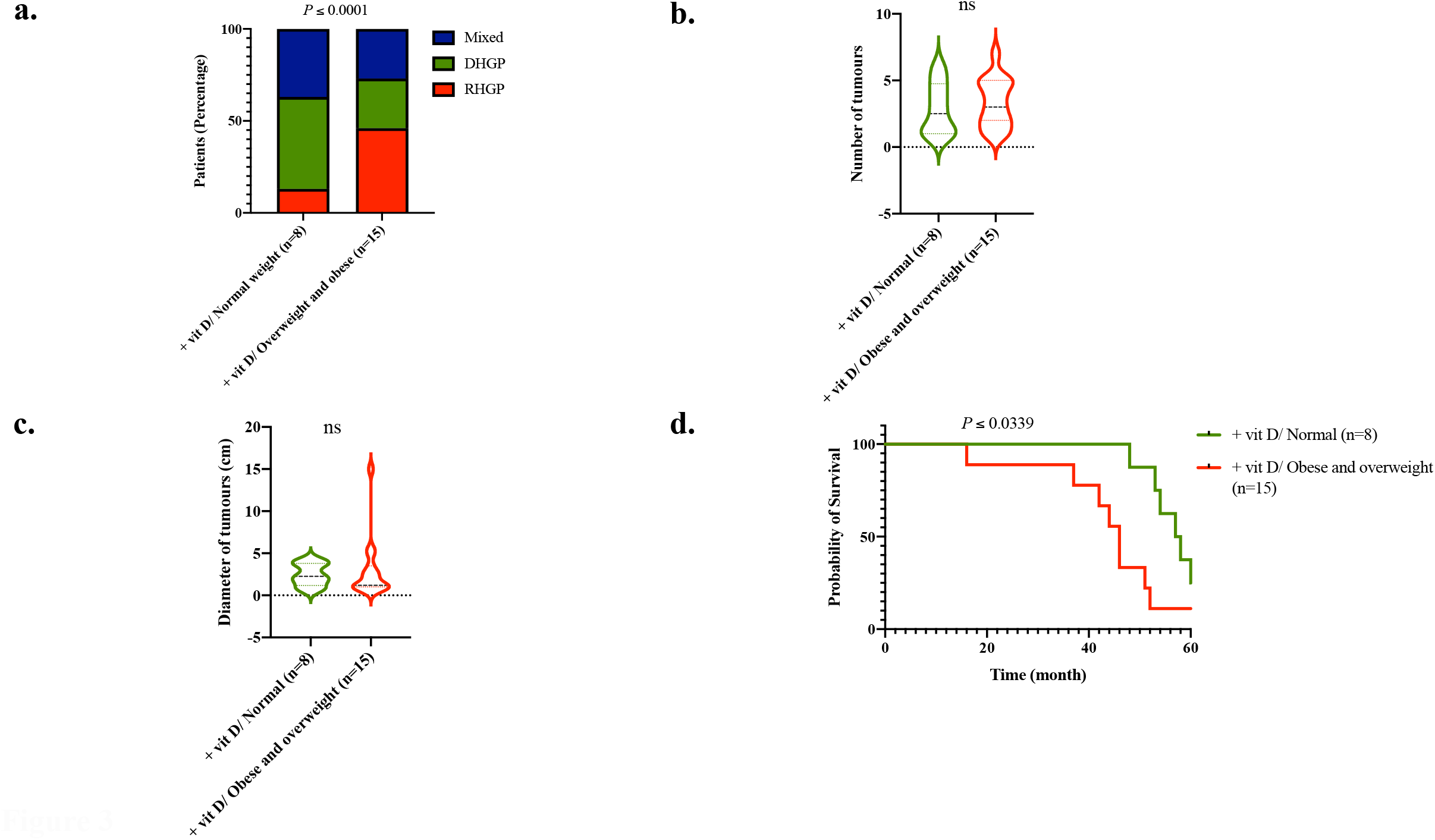
Body mass index of the patient influence the function of vitamin D in CRCLM. **a**. Represents the association between vitamin D supplementations and the HGP of the tumours in CRCLM patients with normal or overweight/obese. **b and c**. Show the number and diameter of CRCLM tumours in either normal or overweight/obese patients that administrated vitamin D supplementation, respectively. **d**. Kaplan–Meier survival analysis shows 5-year OS in the normal or overweight/obese CRCLM patients upon vitamin D supplementation.

### Exposure to vitamin D attenuates EMT and motility of the cancer cells in vitro

We previously showed the association between vessel co-option tumour development and upregulated cancer cell motility^27,29^. Accordingly, high levels of cancer cell motility are mediated by the upregulation of ARP2/3 and EMT markers including vimentin^29^. To understand the molecular mechanisms by which vitamin D attenuate the development of vessel co-option, we examined the effect of vitamin D on co-cultured HT29 colorectal cancer cells with IHH hepatocytes. Insert co-culturing model allows the crosstalk between cancer cells and hepatocytes, similar to vessel co-option lesions^29^. Various molecules are involved in cancer cell motility. Among these molecules, upregulation of ARP2/3 and vimentin significantly contribute to the motility of cancer cells^11-13,29^. As shown in Figure 4, the expression of ARP2/3 and vimentin was upregulated in the co-cultured cancer cells. However, the presence of vitamin D dramatically downregulated both proteins, indicating that vitamin D disrupts the crosstalk between co-cultured cancer cells and hepatocytes, which results in reducing the motility of the cancer cells. However, further investigations are required to determine the molecules that mediate the function of vitamin D. Altogether, these data suggest that vitamin D attenuates the development of vessel co-option tumours in CRCLM via downregulating cancer cell motility.

**Figure 4.**
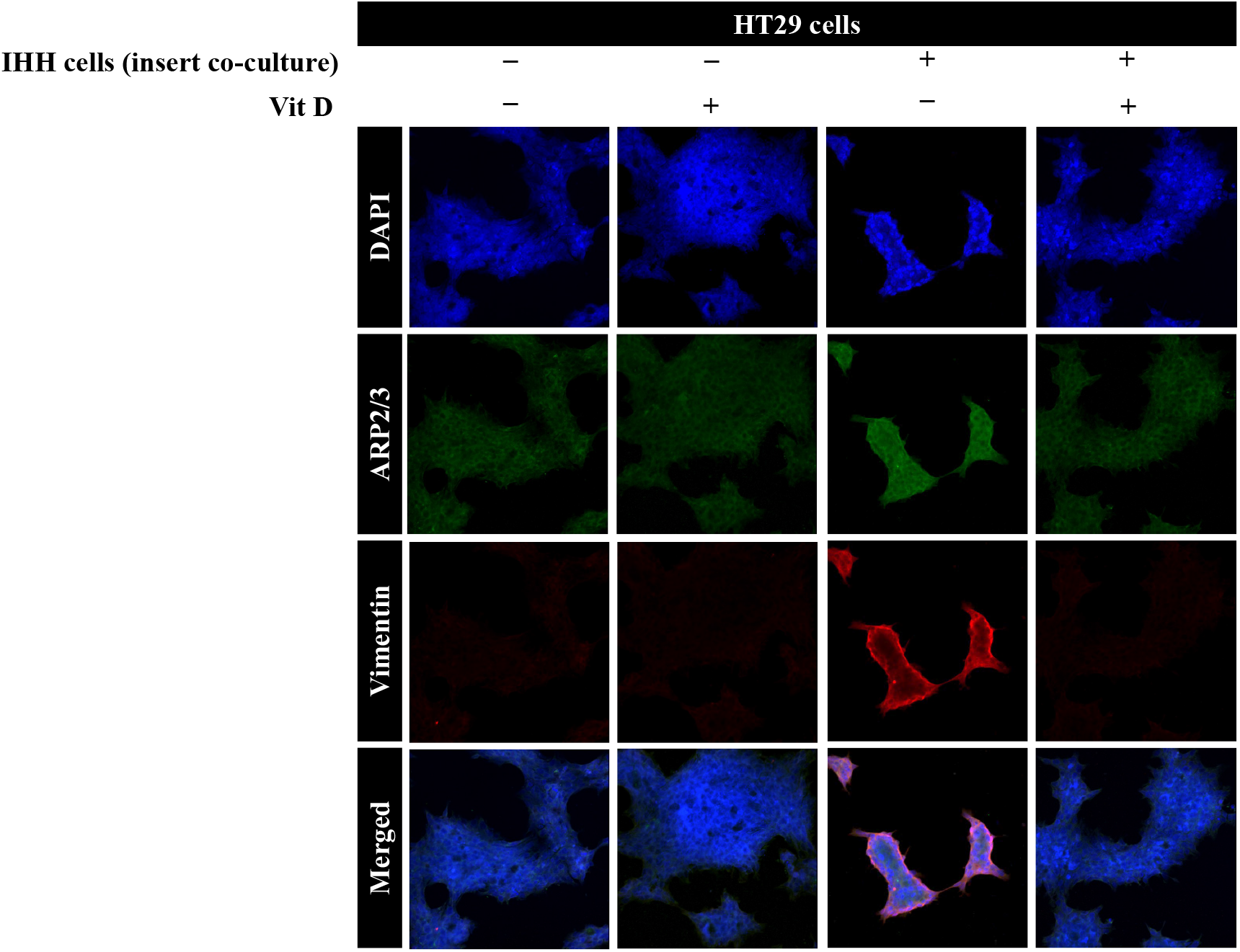
Exposure to vitamin D downregulates the motility and EMT biomarkers in cancer cells in vitro. Immunofluorescence staining showing the expression of motility, ARP2/3, and EMT, vimentin, markers in HT29 colorectal cancer cells upon co-culturing with IHH hepatocytes and/or exposure to vitamin D.

## Discussion

The RHGP tumours are characterized by vessel co-option vascularization and the patients with RHGP tumours have the worse prognosis compared to their counterparts as demonstrated in this manuscript and previous publications^27,39^. Since RHGP tumours do not utilize angiogenesis, the tumours are resistant to anti-angiogenic agents, such as Bevacizumab^15,27^. Moreover, vessel co-option tumours are characterized by resistance against chemotherapy^8,15,27^. Thus, increasing the development of angiogenic tumours or converting the vessel co-option tumours to angiogenic tumours significantly improves CRCLM response to anti-angiogenic agents and chemoresistance, subsequently improving the prognosis of CRCLM patients.

The role of vitamin D supplementation has been investigated in different cancers^30–33^. However, its role in CRCLM is poorly understood. In this manuscript, we found a positive correlation between vitamin D supplementation and the prognosis of CRCLM patients. The positive effect of vitamin D supplementation in CRCLM is likely incited by its role in the attenuation of vessel co-option development, which is mediated by cancer cell motility and EMT^29^. Vitamin D driven inhibition of cancer cell motility and migration has been shown previously. In this context, Liu et al.^40^ have demonstrated that vitamin D inhibits cancer cell migration in lung cancer. Similar effect of vitamin D has been reported in other types of cancers, such as thyroid^30^, prostate, breast, gastric^41^, and colorectal^42^ cancer. Moreover, vitamin D has been shown to suppress cancer cell proliferation and induce apoptosis^40^. Therefore, further investigations are required to identify the effect of vitamin D in cancer cell proliferation, as well as response to anti-cancer therapies in CRCLM.

The role of vitamin D in vascularization is conflicting. Vitamin D has been shown to increase angiogenesis by reducing the activation, proliferation, migration, and germination of endothelial cells^21^. Our clinical data also suggested a positive association between vitamin D supplementation and the presence of angiogenic tumours in CRCLM patients. However, various studies reported the inhibition of tumour angiogenesis upon vitamin D supplementation^22,23^.

## Conclusions

Our data demonstrated a new function of vitamin D in cancer, and its administration may provide promising new opportunities for treating CRCLM patients with vessel co-option tumours. The role of vitamin D in CRCLM response to anti-angiogenic agents and chemotherapy warrants further investigation.

## Data Availability

All data produced in the present study are available upon reasonable request to the authors

## Data Availability

All data produced in the present study are available upon reasonable request to the authors

## Author Contributions

M.R., A.L. and P.M. co-conceived the study. L.K. and S.P. collected the clinical data. M.R. analyzed the clinical data and performed cell culturing, immunofluorescence, data curation, writing and preparation of the manuscript. A.K.L. and D.K. assisted in cell culturing. P.M. review the manuscript and funding acquisition.

## Institutional Review Board Statement

The study was approved by the McGill University Health Centre Institutional Review Board.

## Data Availability Statement

All data produced in the present study are available upon reasonable request to the authors

## Acknowledgments

The authors would like to acknowledge the support provided by Dana Massaro and Ken Verdoni Liver Metastases Research Fellowship.

## References

1. Rawla, P., Sunkara, T. & Barsouk, A. Epidemiology of colorectal cancer: Incidence, mortality, survival, and risk factors. Prz. Gastroenterol. 14, 89–103 (2019).

2. Siegel, R. L., Miller, K. D., Fuchs, H. E. & Jemal, A. Cancer statistics, 2022. CA. Cancer J. Clin. 72, 7–33 (2022).

3. Pan, Z. et al. Is there a survival benefit from adjuvant chemotherapy for patients with liver oligometastases from colorectal cancer after curative resection? Cancer Commun. 38, 1–10 (2018).

4. House, M. G. et al. Survival after Hepatic Resection for Metastatic Colorectal Cancer: Trends in Outcomes for 1,600 Patients during Two Decades at a Single Institution. J. Am. Coll. Surg. 210, 744–752 (2010).

5. Guadagni, S. et al. Real-life multidisciplinary treatment for unresectable colorectal cancer liver metastases including hepatic artery infusion with chemo-filtration and liquid biopsy precision oncotherapy: observational cohort study. J. Cancer Res. Clin. Oncol. 146, 1273–1290 (2020).

6. Fusai, G. & Davidson, B. R. Strategies to Increase the Resectability of Liver Metastases from Colorectal Cancer. Dig. Surg. 20, 481–496 (2003).

7. Rocha, F. G. & Helton, W. S. Resectability of colorectal liver metastases : an evolving definition. HPB 14, 283–284 (2012).

8. Frentzas, S., Lum, C. & Chen, T.-Y. Angiogenesis and Its Role in the Tumour Microenvironment: A Target for Cancer Therapy. in Current Cancer Treatment 1–14 (2019). doi:10.5772/intechopen.89667

9. Rada, M. et al. Cancer cells induce hepatocytes apoptosis in co-opted colorectal cancer liver metastatic lesions. bioRxiv 429243, (2021).

10. Rada, M., Hassan, N., Lazaris, A. & Metrakos, P. The molecular mechanisms underlying neutrophil infiltration in vessel co-opting colorectal cancer liver metastases. Front. Oncol. (2022). doi:10.3389/fonc.2022.1004793

11. Rada, M., Lazaris, A., Kapelanski-Lamoureux, A., Mayer, T. Z. & Metrakos, P. Tumor microenvironment conditions that favor vessel co-option in colorectal cancer liver metastases: A theoretical model. Semin. Cancer Biol. 71, 52–64 (2021).

12. Rada, M. et al. Angiopoietin-1 Upregulates Cancer Cell Motility in Colorectal Cancer Liver Metastases through Actin-Related Protein 2/3. Cancers (Basel). 14, 2540 (2022).

13. Rada, M. et al. Cancer Cells Promote Phenotypic Alterations in Hepatocytes at the Edge of Cancer Cell Nests to Facilitate Vessel Co-Option Establishment in Colorectal Cancer Liver Metastases. 1–19 (2022).

14. Palmieri, V. et al. Neutrophils expressing lysyl oxidase-like 4 protein are present in colorectal cancer liver metastases resistant to anti-angiogenic therapy. J. Pathol. 251, 213–223 (2020).

15. Kuczynski, E. A., Vermeulen, P. B., Pezzella, F., Kerbel, R. S. & Reynolds, A. R. Vessel co-option in cancer. Nat. Rev. Clin. Oncol. 16, 469–493 (2019).

16. Shaw, E., Massaro, N. & Brockton, N. T. The role of vitamin D in hepatic metastases from colorectal cancer. Clin. Transl. Oncol. 20, 259–273 (2018).

17. Gupta, D., Vashi, P. G., Trukova, K., Lis, C. G. & Lammersfeld, C. A. Prevalence of serum vitamin D deficiency and insufficiency in cancer: Review of the epidemiological literature. Exp. Ther. Med. 2, 181–193 (2011).

18. Negri, M. et al. Vitamin D-induced molecular mechanisms to potentiate cancer therapy and to reverse drug-resistance in cancer cells. Nutrients 12, 1–25 (2020).

19. Chandler, P. D. et al. Effect of Vitamin D3Supplements on Development of Advanced Cancer: A Secondary Analysis of the VITAL Randomized Clinical Trial. JAMA Netw. Open 3, (2020).

20. Ibrahim, N. S. et al. Angiopoietin1 deficiency in hepatocytes affects the growth of colorectal cancer liver metastases (Crclm). Cancers (Basel). 12, (2020).

21. Aliashrafi, S. & Ebrahimi-Mameghan, M. A systematic review on vitamin D and angiogenesis. in Angiogenesis 7, 1–78 (2017).

22. Chakraborti, C. Vitamin D as a promising anticancer agent. Indian J. Pharmacol. 43, 113–120 (2011).

23. Mantell, D. J., Owens, P. E., Bundred, N. J., Mawer, E. B. & Canfield, A. E. 1α,25-Dihydroxyvitamin D3 inhibits angiogenesis in vitro and in vivo. Circ. Res. 87, 214–220 (2000).

24. Dehghani, L. et al. Can vitamin D suppress endothelial cells apoptosis in multiple sclerosis patients? Int. J. Prev. Med. 4, S211–S215 (2013).

25. Gupta, M. K. & Qin, R. Y. Mechanism and its regulation of tumor-induced angiogenesis. World J. Gastroenterol. 9, 1144–1155 (2003).

26. Grundmann, M. et al. Vitamin D improves the angiogenic properties of endothelial progenitor cells. Am. J. Physiol. - Cell Physiol. 303, 954–962 (2012).

27. Frentzas, S. et al. Vessel co-option mediates resistance to anti-angiogenic therapy in liver metastases. Nat. Med. 22, 1294–1302 (2016).

28. Rada, M. et al. Disruption of integrin alpha-5/beta-1-dependent transforming growth factor beta-1 signaling pathway attenuates vessel co-option in colorectal cancer liver metastases. bioRxiv 2003–2005 (2022). doi:10.1101/2022.05.24.493291

29. Rada, M. et al. Runt related transcription factor-1 plays a central role in vessel co-option of colorectal cancer liver metastases. Commun. Biol. 4, 1–15 (2021).

30. Coperchini, F. et al. Vitamin D Reduces Thyroid Cancer Cells Migration Independently From the Modulation of CCL2 and CXCL8 Chemokines Secretion. Front. Endocrinol. (Lausanne). 13, 1–7 (2022).

31. Blasiak, J., Chojnacki, J., Pawlowska, E., Jablkowska, A. & Chojnacki, C. Vitamin D May Protect against Breast Cancer through the Regulation of Long Noncoding RNAs by VDR Signaling. Int. J. Mol. Sci. 23, (2022).

32. Wang, L., Zhou, S. & Guo, B. Vitamin D suppresses ovarian cancer growth and invasion by targeting long non-coding RNA CCAT2. Int. J. Mol. Sci. 21, (2020).

33. Bintintan, V. V. Vitamin D as a Potential Therapeutic Target and Prognostic Marker for Colorectal Cancer. EBioMedicine 31, 11–12 (2018).

34. Bajbouj, K. et al. Vitamin D Exerts Significant Antitumor Effects by Suppressing Vasculogenic Mimicry in Breast Cancer Cells. Front. Oncol. 12, 1–12 (2022).

35. Palmieri, V. et al. Neutrophils expressing lysyl oxidase-like 4 protein are present in colorectal cancer liver metastases resistant to anti-angiogenic therapy. J. Pathol. 251, 213–223 (2020).

36. Ramdass, V. et al. Association Between Obesity and Cancer Mortality: An Internal Medicine Outpatient Clinic Perspective. J. Clin. Med. Res. 13, 377–386 (2021).

37. Saleh, K. et al. Impact of body mass index on overall survival in patients with metastatic breast cancer. Breast 55, 16–24 (2021).

38. Chiu, C. C. et al. Correlation of body mass index with oncologic outcomes in colorectal cancer patients: A large population-based study. Cancers (Basel). 13, 1–19 (2021).

39. Rada, M., Krzywon, L., Kapelanski-lamoureux, A. & Petrillo, S. High levels of serum cholesterol positively correlate with the risk of the development of vessel co-opting tumours in colorectal cancer liver metastases. medRxiv 1–25 (2022).

40. Liu, N. et al. Inhibition of lung cancer by vitamin D depends on downregulation of histidine-rich calcium-binding protein. J. Adv. Res. 29, 13–22 (2021).

41. Jeon, S. M. & Shin, E. A. Exploring vitamin D metabolism and function in cancer. Exp. Mol. Med. 50, (2018).

42. Zhu, Y. et al. MEG3 Activated by Vitamin D Inhibits Colorectal Cancer Cells Proliferation and Migration via Regulating Clusterin. EBioMedicine 30, 148–157 (2018).

